# Repurposing Clinical MRI Archives for Multiple Sclerosis Research with a Flexible, Single-Modality Approach: New Insights from Old Scans

**DOI:** 10.1101/2024.03.29.24305083

**Authors:** Philipp Goebl, Jed Wingrove, Omar Abdelmannan, Barbara Brito Vega, Jonathan Stutters, Silvia Da Graca Ramos, Owain Kenway, Thomas Rosoor, Evangeline Wassmer, Jeremy Chataway, Douglas Arnold, Louis Collins, Cheryl Hemmingway, Sridar Narayanan, Declan Chard, Juan Eugenio Iglesias, Frederik Barkhof, Yael Hacohen, Alan Thompson, Daniel Alexander, Olga Ciccarelli, Arman Eshaghi

## Abstract

In multiple sclerosis (MS), magnetic resonance imaging (MRI) biomarkers are critical for research in diagnosis, prognosis and assessing treatment efficacy. Traditionally, extracting relevant biomarkers of disease activity and neurodegeneration requires multimodal MRI protocols, limiting the use of the already existing vast amount of incomplete or single-modality MRI data which are acquired in clinical settings. We developed MindGlide, a deep learning model that extracts volums of brain regions and lesion from a single MRI modality, simplifying analysis and enabling the use of heterogeneous clinical archives. We trained MindGlide on a dataset of 4,247 brain MRI scans from 2,934 MS patients across 592 MRI scanners and validated it on 14,952 brain MRI scans from 1001 patients from three unseen external validation cohorts including 161 adolescent patients. Using dice scores, we demonstrated that MindGlide accurately estimated white matter lesion, cortical, and deep grey matter volumes. These volumes correlated with disability (Expanded Disability Status Scale, absolute correlation coefficients 0.1-0.2, p<0.05), and MindGlide outperformed an established tool in this regard. MindGlide robustly detected treatment effects across clinical trials, including disease activity and neurodegeneration (as shown by lesion accrual and brain tissue loss, respectively), even when analysing MRI modalities not traditionally used for such detailed measurements. Our results indicate the potential to indirectly reduce scan time and drug development costs in clinical trials while directly transforming the utility of retrospective analysis of real-world data acquired in clinical settings. As a consequence, scan time will be reduced and, in turn, the cost of trials.

## INTRODUCTION

Multiple sclerosis (MS) is a chronic disabling condition affecting over 2.5 million people worldwide, with a disproportionate impact on young populations^1^. MRI-based biomarkers are central to MS clinical trials, where they represent primary and secondary efficacy endpoints. Specialised imaging protocols include multiple sequences to capture distinct aspects of disease evolution: new or enlarging lesions indicate active inflammation, while brain atrophy is a proxy for neurodegeneration. However, these comprehensive protocols are time-consuming and costly ^2,3^. Simplifying MRI analysis, particularly by enabling biomarker extraction from a single modality, can accelerate drug development by making clinical trials cheaper and expand research possibilities by offering the opportunity to analyse MRI scans previously acquired according to standard-of-care clinical protocols.

While MRI monitoring of treatment response in routine care focuses on new or enlarging lesions ^4^, other disease processes such as brain atrophy (even without overt disease activity) contribute to MS worsening^5,6^. Unlike the standardised protocols used in experimental clinical trials, the variability of archival routine clinical scans presents a major obstacle to the reliable, automatic quantification of brain structures and lesions. We aimed to develop a computationally efficient tool to process these highly heterogeneous scans across modalities and qualities. Such a tool would enable the extraction of key MRI biomarkers in diverse clinical settings, paving the way for personalised medicine algorithms. Furthermore, it can help to overcome a bias of past MS trials not reflecting populational diversity and instead being comprised of predominantly Caucasian participants. Using routine care clinical MRI images instead of clinical trial MRI images, such a tool opens the option to better include the whole spectrum of a society and analyse potential differences across biomarkers.

Recent advances in deep learning and generative models have enhanced our ability to analyse real-world medical images. Tools like SynthSeg and others exemplify this progress, primarily focusing on brain segmentation via lesion inpainting^7–12^. SAMSEG-lesion is a newly introduced model that robustly segments lesions and brain structures across MRI modalities^10^. A model named WMH-SynthSeg has demonstrated the potential for segmenting white matter hyperintensities and brain anatomy from scans of any resolution and contrast, including low-field portable MRI^13^. However, existing tools have not been developed or validated for MS clinical trials or highly heterogeneous real-world MS clinical data. This highlights the urgent need for solutions that extract MRI biomarkers, including lesion load and changes in brain volume, from the often single-modality scans acquired in routine care, including T1-weighted, T2-weighted and FLAIR images, and indirectly accelerate drug development by reducing scan time.

Here, we present MindGlide, a deep-learning model that addresses these limitations. We designed MindGlide to (1) efficiently (in less than a minute with no pre-processing required by the user) quantify brain structures and lesions. It does so in a modality-agnostic way from a single MRI sequence input. Furthermore, Mindglide can (2) measure treatment effects using MRI sequences not typically analysed for these purposes, and (3) demonstrate the potential of routine MRI data to detect new lesions and subtle brain tissue loss, even when the ideal imaging sequences are unavailable.

## Methods

### Overview

We developed MindGlide a 3D convolutional neural network (CNN) to segment specific brain structures and white matter lesions. We designed it to process brain MRI robustly across commonly available modalities in hospital archives. It handles the variation in tissue intensities caused by diverse MRI acquisition parameters (T1-weighted, T2-weighted, Proton Density (PD), and T2-Fluid Attenuated Inversion Recovery [FLAIR]), including both 2D (two-dimensional) and 3D (three-dimensional) scans. Beyond handling the technical variability of MRI scans, MindGlide also effectively addresses different forms of disease progression that occur across different age groups.

### Study Design and Participants

We used distinct training and external validation datasets consisting exclusively of patients with MS. **Table 1** shows patient characteristics in the training and external validation sets. The training set comprised seven previously published clinical trials^14–22^ and one observational cohort (Supplemental Table 1). This set included only T1-weighted and FLAIR MRI modalities and synthetically generated scans (see below for details). We froze model parameters after training completion.

**Table 1.**
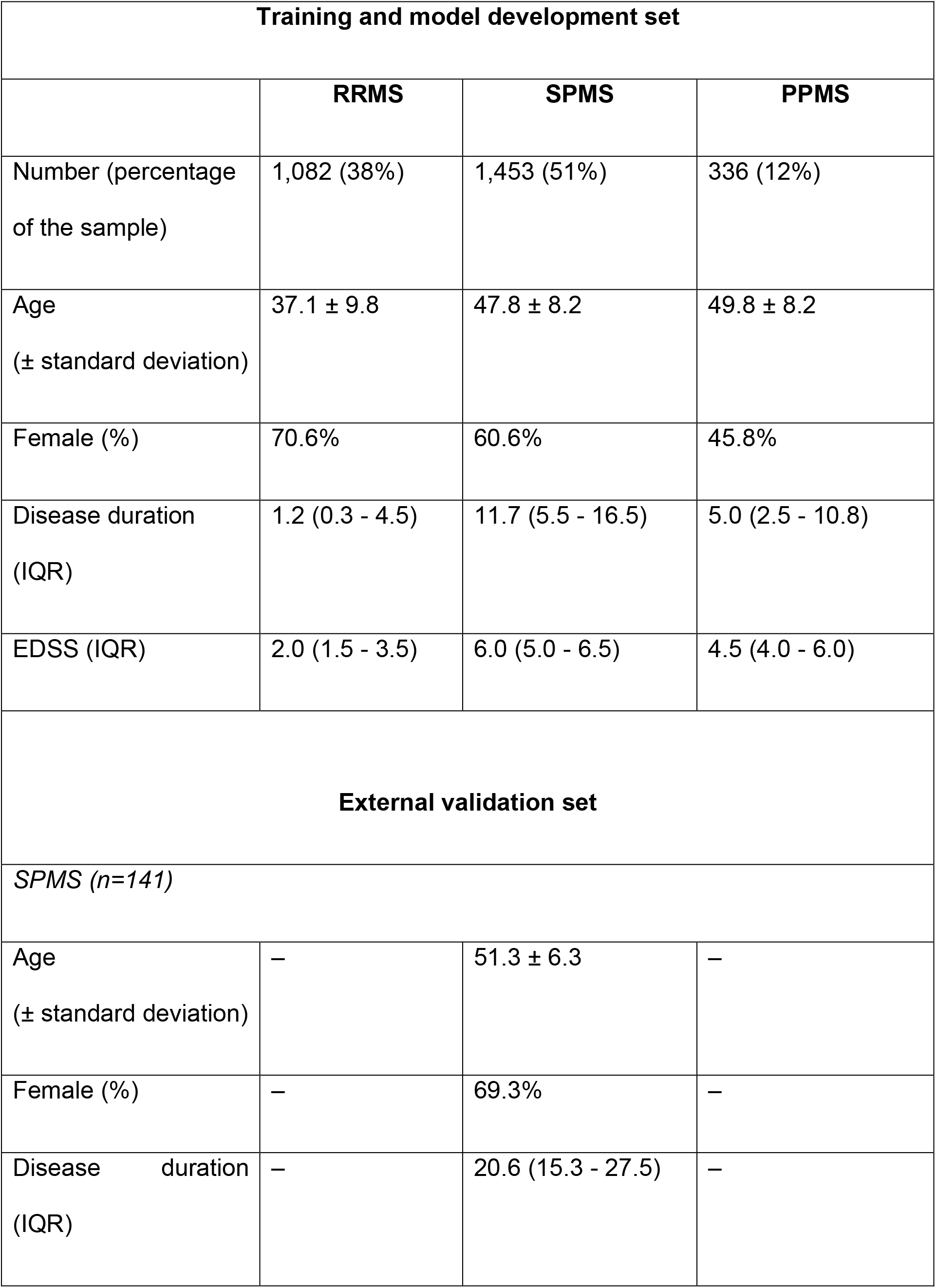
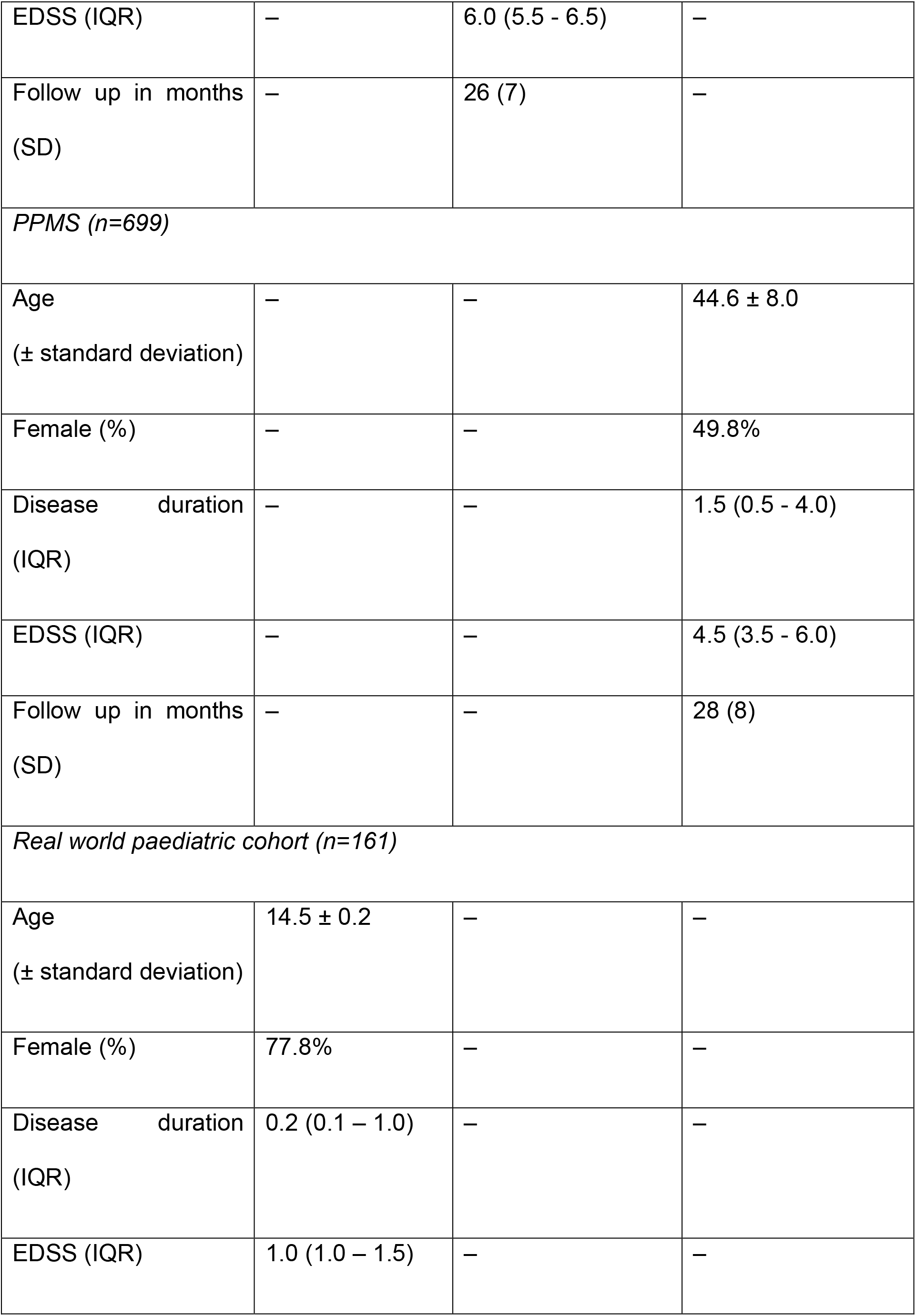
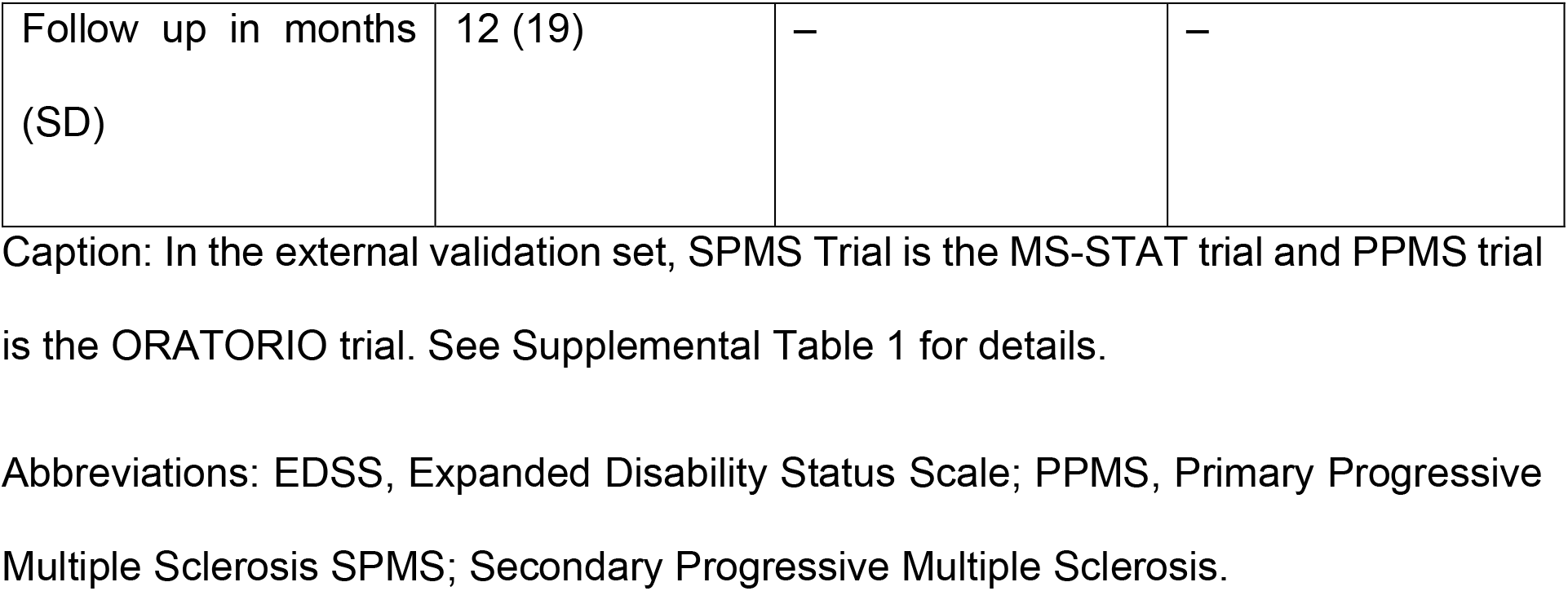
Patient characteristics in the training and external validation sets.

| Training and model development set |  |  |  |
| --- | --- | --- | --- |
|  | RRMS | SPMS | PPMS |
| Number (percentage of the sample) | 1,082 (38%) | 1,453 (51%) | 336 (12%) |
| Age<br>(± standard deviation) | 37.1 ± 9.8 | 47.8 ± 8.2 | 49.8 ± 8.2 |
| Female (%) | 70.6% | 60.6% | 45.8% |
| Disease duration<br>(IQR) | 1.2 (0.3 - 4.5) | 11.7 (5.5 - 16.5) | 5.0 (2.5 - 10.8) |
| EDSS (IQR) | 2.0 (1.5 - 3.5) | 6.0 (5.0 - 6.5) | 4.5 (4.0 - 6.0) |
| External validation set |  |  |  |
| *SPMS (n=141)* |  |  |  |
| Age<br>(± standard deviation) | – | 51.3 ± 6.3 | – |
| Female (%) | – | 69.3% | – |
| Disease duration<br>(IQR) | – | 20.6 (15.3 - 27.5) | – |
| EDSS (IQR) | – | 6.0 (5.5 - 6.5) | – |
| Follow up in months (SD) | – | 26 (7) | – |
| *PPMS (n=699)* |  |  |  |
| Age (± standard deviation) | – | – | 44.6 ± 8.0 |
| Female (%) | – | – | 49.8% |
| Disease duration (IQR) | – | – | 1.5 (0.5 - 4.0) |
| EDSS (IQR) | – | – | 4.5 (3.5 - 6.0) |
| Follow up in months (SD) | – | – | 28 (8) |
| *Real world paediatric cohort (n=161)* |  |  |  |
| Age (± standard deviation) | 14.5 ± 0.2 | – | – |
| Female (%) | 77.8% | – | – |
| Disease duration (IQR) | 0.2 (0.1 – 1.0) | – | – |
| EDSS (IQR) | 1.0 (1.0 – 1.5) | – | – |
| Follow up in months<br>(SD) | 12 (19) | – | – |

Caption: In the external validation set, SPMS Trial is the MS-STAT trial and PPMS trial is the ORATORIO trial. See Supplemental Table 1 for details.
Abbreviations: EDSS, Expanded Disability Status Scale; PPMS, Primary Progressive Multiple Sclerosis SPMS; Secondary Progressive Multiple Sclerosis.

To test MindGlide’s generalisability, we employed an external validation set of two clinical trials^2,23^ and a real-world cohort of paediatric MS patients. This set encompassed unseen T2-weighted and PD MRI modalities in addition to T1-weighted and FLAIR scans. We specifically selected the paediatric cohort to test robustness across age groups, given the typically more inflammatory disease course and larger lesion volume relative to skull size in early-onset cases.

### Ethical approval

This study received ethical approval from the Institutional Review Board under the auspices of the International Progressive Multiple Sclerosis Alliance (www.progressivemsalliance.org) at the Montreal Neurological Institute, Canada (IRB00010120) and by Great Ormond Street Hospital Research and Development Department (reference: 16NC10).

### Model development

We developed MindGlide, using “nnU-Net”, a 3D CNN building on the widespread U-Net architecture.^24^ nnU-Net yields state-of-the-art results (e.g., it has won several recent challenges^12,25^) while featuring automatic self-configuration – and thus bypassing the costly hyperparameter tuning procedure. We trained MindGlide to segment brain regions and lesions in MS patients, accommodating real-world MRI variations and artefacts that often hinder traditional image processing software. Our primary goal was to ensure generalisation across MRI modalities with minimal or no pre-processing at inference, even for modalities unseen during training (PD and T2-weighted). This aligns with successful approaches in other brain imaging studies^7,8^. **Figure 1** illustrates our training and external validation strategy and the associated steps.

**Figure 1.**
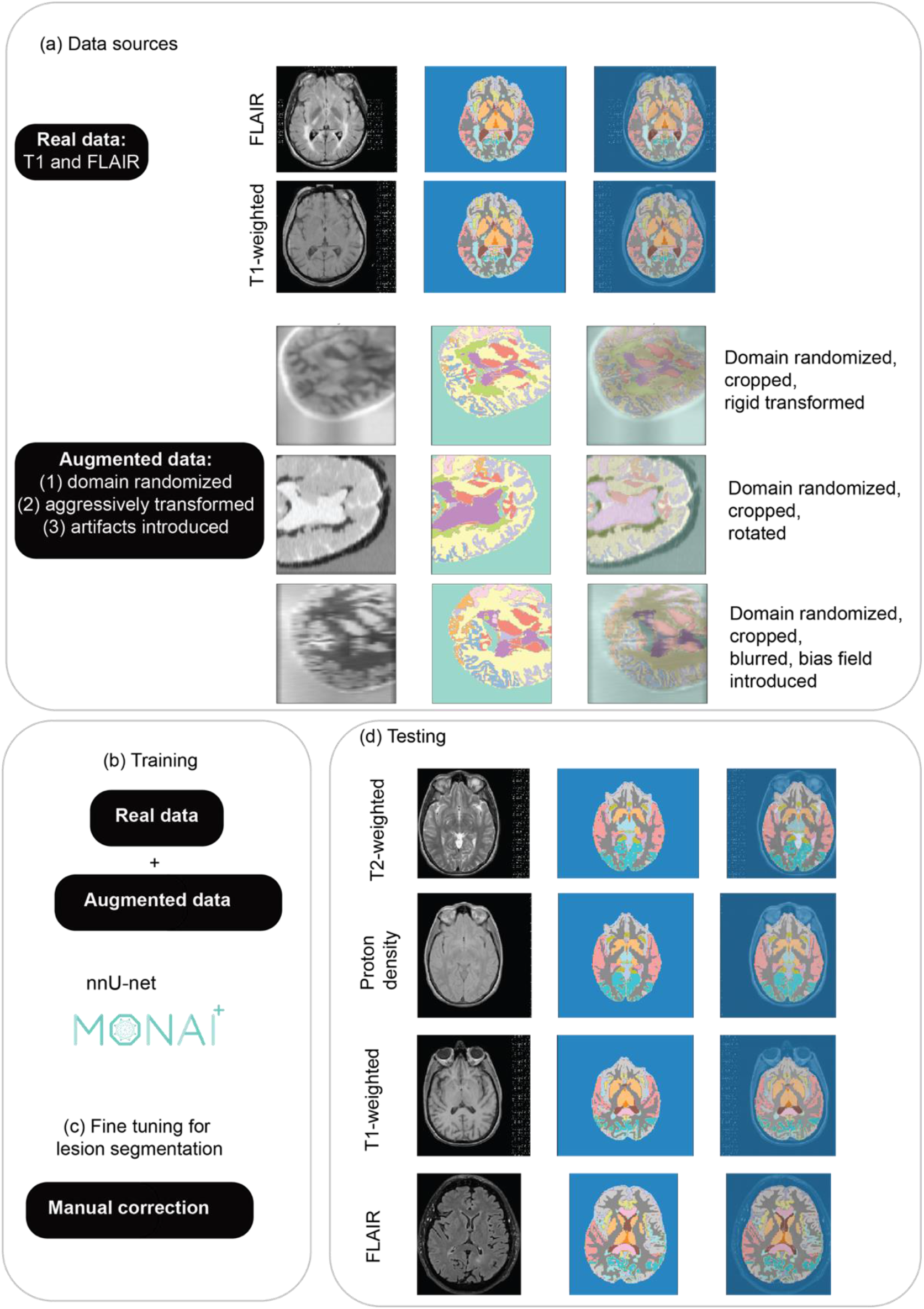
Developing and testing the MindGlide model. MindGlide model enables highly efficient and robust MRI segmentation. Segmenting and quantifying lesions on heterogeneous modalities with minimal pre-processing (and no pre-processing required by the user). MindGlide model generalizes to tasks not used to train the model, such as segmenting T2-weighted and PD MRI scans in unseen data sets. Abbreviations: FLAIR, Fluid Attenuated Inversion Recovery.

#### Generating training labels

Supervised models like MindGlide require large, accurately labelled datasets for robust performance. Manually creating such labels, considered the ’gold standard’, is time-consuming and impractical for diverse real-world data, especially when the quality of scans may hinder manual labelling. Therefore, we leveraged existing segmentations from our existing datasets of phase two and three clinical trials previously published^26^ with Geodesic Information Flows software (GIF v3.0) as explained in our previous publication^27^, with additional manual quality control. These labels, derived from the Neuromorphometrics atlas (http://neuromorphometrics.com), were grouped into 18 regions (**Supplemental Table 2**). To reduce the number of labels and make MindGlide more comparable to other brain image segmentation tools we performed this label grouping according to the hierarchical model of the Mindboggle project (https://mindboggle.info/braincolor). We used a validated lesion segmentation model ^26,28^, to incorporate lesion data, creating a single file with 20 labels (18 brain regions, 1 lesion, 1 background) for training. We merged the regional maps with the lesion map to have one file with 20 labels for training our model (18 brain regions, 1 lesion label, and 1 background label). We employed existing expert-labelled ’ground truth’ data for external validation in a smaller number of individuals, as described below.

#### Image Pre-processing for Model Training

We employed a minimal pre-processing pipeline. We first standardised image resolution to 1.0mm isotropic voxels, ensuring consistency across datasets per the nnU-Net design^12,25^. We then extracted 128x128x64 voxel patches using a sliding window technique, to optimise memory and computational efficiency during training. While we used data augmentation during training (see below), no further pre-processing was performed at inference.

#### Image Augmentation and synthetic data

Data augmentation is systematic random modification of training data in order to artificially extend the support of its statistical distribution. It is crucial for model generalisability and preventing overfitting. To minimise post-training pre-processing and broaden MindGlide’s adaptability, we used two techniques: (1) distorting real scans in the spaces of geometry and image intensities and (2) generating synthetic ones. Synthetic data generation offers greater flexibility than mere distortion. We employed domain randomisation (Figure 1a), resulting in intensity variations that prepared the model for diverse MRI modalities. As shown in Figure 1d, we performed image augmentation with T1-weighted and FLAIR scans during training.

We used SynthSeg version 2.0 for synthetic data generation and MONAI version 1.2.0 for augmentation during training ^7,9,11,29^. We generated synthetic scans of varying contrasts directly from the training dataset’s labels (units are as defined by the software)^9^:

- Left-right flipping (0.5 probability).
- Scaling (uniform distribution, bounds: 0.85 to 1.15).
- Rotation (uniform distribution, bounds: -15 to 15 degrees).
- Elastic deformation (scale: 0.04, standard deviation: 1).
- Bias field corruption (scale: 0.25, standard deviation: 0.5).
- Random low-resolution resampling (uniform distribution, 1 to 9 mm per dimension).
- Domain randomisation^9^ (varying voxel intensities of synthetic scans per tissue class) **Figure 1a** illustrates examples of the synthetic data generated. The model architecture, detailed in the Supplemental Material (Model Architecture), has one input channel (receiving a single MR modality) and 20 output channels (generating 20 labelled segmentations).

### External validation using unseen data

We performed cross-sectional and longitudinal analyses to assess MindGlide’s performance for its validation and reliability in unseen cohorts.

#### Cross-sectional analyses

This involved clinical validation by correlating segmented structure and lesion volumes with Expanded Disability Status Scale (EDSS) scores, comparing results between MindGlide and SAMSEG ^10^. We chose SAMSEG because it is a recently introduced model, publicly available, as part of Freesurfer, and is one of the few models that can segment multiple different modalities. Manual lesion segmentation by expert neuroradiologists is considered the gold standard in MS. We used two open-source lesion segmentation datasets (MS-30 and ISBI)^30,31^ and assessed cross-sectional performance against manual lesion segmentations (consensus in MS-30, expert rater in ISBI) using lesion volume and voxel-wise spatial metrics (e.g., Dice score, a standard metric for image segmentation overlap).

#### Longitudinal validation in unseen cohorts

We evaluated MindGlide’s ability to detect known treatment effects by analysing data from two successful clinical trials: MS-STAT^23^ (placebo vs. simvastatin in secondary progressive MS) and ORATORIO^2^ (ocrelizumab vs. placebo in primary progressive MS). We calculated the intra-class correlation coefficient (ICC) for percentage brain volume change with MindGlide segmentations and SIENA algorithm^32^ (PBVC), a key trial outcome measure, across MRI modalities (FLAIR, T2-weighted, T1-weighted, and PD in the PPMS trial and T1 and T2-weighted MRI in the SPMS trial).

Additionally, we used a real-world dataset of paediatric relapsing-remitting MS patients from three UK hospitals (Great Ormond Street Hospital, Evelina London Children’s Hospital and Birmingham Children’s Hospital) to study the longitudinal evolution of lesions and brain structures based on available MRI modalities (T1-weighted, T2-weighted and FLAIR). We excluded scans from participants whose FLAIR image slice thickness differed by more than a factor of three across follow-up scans. We did not exclude T1-weighted and T2-weighted MRIs because their slice thicknesses varied by less than a factor of three across visits. All participants in this dataset received a disease-modifying treatment (high efficacy and moderate efficacy treatments). High efficacy treatments included ocrelizumab, natalizumab, rituximab or cladribine and moderate efficacy treatment included interferon betas, fingolimod, dimethyl fumarate or teriflunomide.

#### Reliability analysis in unseen cohorts

Lesion Segmentation across Software: We compared lesion segmentations produced by MindGlide and SAMSEG against ground truth labels (hand-labelled segmentations).

Segmentation Consistency across Modalities: Focusing on the PPMS trial, the only dataset with PD, T2, T1, and FLAIR modalities, we assessed the agreement of segmentations for the same brain structures across these modalities. We used a hierarchical intraclass correlation coefficient (ICC) to account for the fact that these measurements were taken from the same individuals, which introduces inherent correlation (ICC 3).

We measured longitudinal reliability using the ISBI dataset, calculating Intraclass Correlation Coefficients (ICC) between raters and MindGlide. See **Supplemental Material** for details.

#### External validation using manual lesion segmentations

We evaluated MindGlide’s lesion segmentation performance using two openly available datasets with manual lesion masks as ground truth. Cross-sectionally, we assessed lesion load and voxel-wise spatial metrics on 50 FLAIR images from 35 patients, and longitudinally, we calculated the intraclass coefficient (ICC) between raters and MindGlide on the ISBI dataset. For more details, please refer to the Supplemental Material.

### Statistical analysis

We used R version 4.3.0 for all analyses. In cross-sectional analysis, we assessed correlations between segmented brain volumes and EDSS using Spearman’s rank correlation and Fisher Z scores because EDSS is an ordinal variable.

For treatment effect analysis, we employed linear mixed-effects models. Each regional volume or lesion load was the dependent variable in a separate model. Fixed independent variables included time, treatment group, their interaction (time x treatment group), and intracerebral volume (ICV). Random effects, nested by visit within participant ID, accounted for repeated measures and within-subject variability. We did not adjust for other variables because the comparisons were made in data from randomised controlled trials in treatment and control arms. In the real-world data we did not adjust for age (all participants were in their adolescence) and used the same fixed independent variables and random effects. In real-world paediatric dataset, we did not perform a head-to-head comparison of moderate versus high efficacy treatment because participants were not randomised and the small number of children with MS did not allow for causal modelling.

We performed ICC with the Pengouin statistical package for Python 3. We used ICC3 because we had a fixed set of “raters” (segmentation of the same structures by different software[MindGlide vs SAMSEG] or from different modalities).

### Code and data availability

The code, trained models, and computational environment (container) for MindGlide are publicly accessible at https://github.com/MS-PINPOINT/mindGlide. Study data cannot be shared due to contractual agreements with pharmaceutical companies controlling data and the confidentiality requirements for real-world hospital data.

## RESULTS

### Patient characteristics

**Table 1** summarises the patient characteristics of our training and validation datasets. For model development, we used 4,247 real MRI scans (2,092 T1-weighted, 2,155 FLAIR) from 2,871 patients with the following subtypes: SPMS (n=1,453), RRMS (n=1,082), PPMS (n=336). These scans were acquired from 592 MRI scanners. **Supplemental Table 1** provides an overview of the trials included in the training data. We generated 4,303 synthetic scans to augment training and trained MindGlide on a combined dataset of 8,550 real and synthetic images.

Our external validation dataset consisted of 1001 patients from 186 MRI scanners, including the PPMS trial (n=699), SPMS trial (n=141), and a real-world paediatric RRMS cohort (n=161).^2,23^ The PPMS dataset comprised 11,015 MRI scans (2,756 T1-weighted, 2,754 T2-weighted, 2,749 FLAIR, 2,756 PD), all with a slice thickness of 3mm (1mmx1mmx3mm). The SPMS dataset included 763 scans (378 T1-weighted, 385 T2-weighted) with varying slice thicknesses (T1: 1mm isotropic, T2: 3mmx1mmx1mm). The real-world cohort consisted of 161 individuals with 1,478 scans (523 T1-weighted, 475 T2-weighted, 480 FLAIR) and diverse slice thicknesses (median: 3.3mm, range: 0.4-9.0mm). The median follow-up time was 28 months (SD: 8 months) in the PPMS dataset, 26 months (SD: 7 months) in the SPMS dataset, and 12 months (SD: 19 months) in the real-world paediatric cohort. In the real-world cohort, 89 received moderate-efficacy, and 72 received high-efficacy treatments.

### Cross-sectional validation in external cohorts: Correlation between Lesion Volume and Clinical Scores

Figure 2 shows the correlations with EDSS. When using FLAIR scans, the correlation coefficient between MindGlide-derived lesion load and EDSS was 0.127 (95% CI: 0.054 to 0.2; P<0.001), compared to 0.009 (95% CI: -0.065 to 0.083; P=0.813) between SAMSEG-derived lesion load and EDSS. When using T2 sequences, MindGlide-derived lesion load showed a correlation coefficient of 0.150 (95% CI: 0.077 to 0.221; P<0.001), versus SAMSEG’s 0.086 (95% CI: 0.012 to 0.159; P=0.022). There was a statistically significant difference between the correlation coefficients when using MindGlide-derived lesion load or SAMSEG-derived lesion load on FLAIR (P=0.026) but not on T2 sequences (P=0.227).

**Figure 2.**
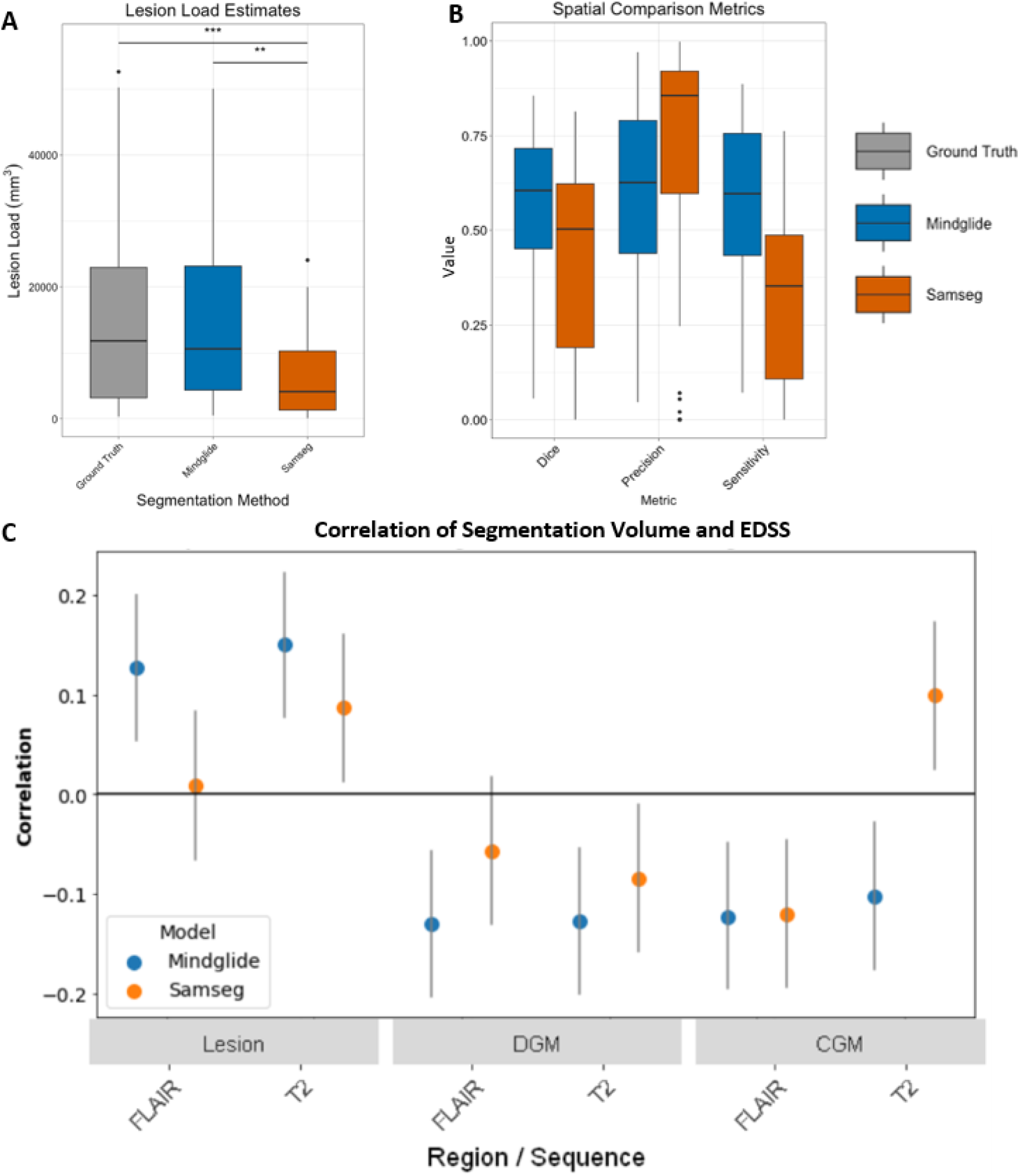
Performance comparisons with other software. (A) Boxplot displaying Lesion Load estimates (mm^3^) and distributions measured using ground truth manual delineations (grey), MindGlide (blue) and Freesurfer’s SAMSEG (orange). Lesion load estimates between Ground truth and SAMSEG and MindGlide and SAMSEG methods were significantly different (paired t-tests). (B) Boxplot displaying Dice scores, Sensitivity and Precision measurements for both MindGlide (blue) and SAMSEG (orange) delineated lesions. *** P < 0.001, ** P < 0.01, n=50. In (C) we calculated Spearman’s correlation coefficients for regional brain volumes obtained from MindGlide and Fressurfer’s SAMSEG against the expanded disability status scale (EDSS). The analysis evaluates the association strength of lesion, deep grey matter (DGM), and cortical grey matter (CGM) volumes with clinical disability, across FLAIR and T2 MRI sequences. MindGlide’s performance is compared to SAMSEG, highlighting which tool’s volumetric assessments have a stronger correlation with the progression of disability as measured by EDSS. For all tested regions and sequences MindGlide’s output shows stronger correlation with EDSS scores. Error bars represent 95% CI. N = 699.

Regarding the correlation between DGM volumes and EDSS scores, MindGlide-derived DGM volume demonstrated a correlation coefficient of -0.130 on FLAIR sequences (95% confidence interval [CI]: -0.202 to -0.057; P<0.001) and -0.128 on T2-weighted sequences (95% CI: -0.200 to -0.054; P<0.001). In contrast, SAMSEG-derived DGM volume yielded correlation coefficients of -0.057 (95% CI: -0.130 to 0.018; P=0.134) and -0.084 (95% CI: -0.157 to -0.010; P=0.026) for FLAIR and T2-weighted sequences, respectively. No statistically significant differences were observed between the correlation coefficients obtained using MindGlide and SAMSEG on either FLAIR (P=0.166) or T2-weighted sequences (P=0.412).

For CGM, MindGlide-derived CGM volume and EDSS correlation coefficients were -0.123 on FLAIR (95% CI: -0.195 to -0.049; P=0.001) and -0.102 on T2 sequences (95% CI: -0.175 to -0.028; P=0.007). SAMSEG-derived CGM volume and EDSS correlation coefficients were -0.121 on FLAIR (95% CI: -0.193 to -0.047; P=0.001) and 0.099 on T2 sequences (95% CI: 0.025 to 0.172; P=0.008). There was a statistically significant difference between the correlation coefficients when using MindGlide or SAMSEG on T2 sequences (P<0.001) but not on FLAIR (P=0.966).

Figure 2b shows the degree of agreement (dice score) of MindGlide-derived volumes and SAMSEG-derived volumes with ground truth hand-labelled lesions on the same scans. The median dice score for MindGlide was 0.606 and for SAMSEG was 0.504. Supplemental Table 3 summarises the cross-software comparison of lesion segmentations (MindGlide and SAMSEG).

### Treatment effects on lesion accrual

When we tested longitudinal lesion accrual in the SPMS trial (simvastatin vs placebo), using T2-weighted MRI, the rate of lesion accrual was, on average, numerically faster in the placebo group than in the treatment group (1.12 mL/year vs 0.768 mL/year; P=0.054).^23^ Using 3D T1-weighted MRI, the rate of hypointense lesion accrual was statistically significantly faster in the placebo group than in the treatment group (1.874 mL/year vs 1.071 mL/year; P=0.005).

As shown in Figure 3, In the PPMS trial (ocrelizumab vs placebo), using T2-weighted MRI, the rate of lesion accrual was faster in the placebo group than in the treatment group (1.103 mL/year vs 0.399 mL/year; P<0.001).^2^ Using FLAIR, the rate of lesion accrual was 1.042 mL/year in the placebo group and 0.141 mL/year in the treatment group (P<0.001). Using the PD MRI modality, the rate of lesion accrual was 0.633 mL/year in the placebo group and 0.91 mL/year in the treatment group (P<0.001). Using T1-weighted MRI, the rate of hypointense lesion accrual was 1.225 mL/year in the placebo group and 0.648 mL/year in the treatment group (P<0.001).

**Figure 3.**
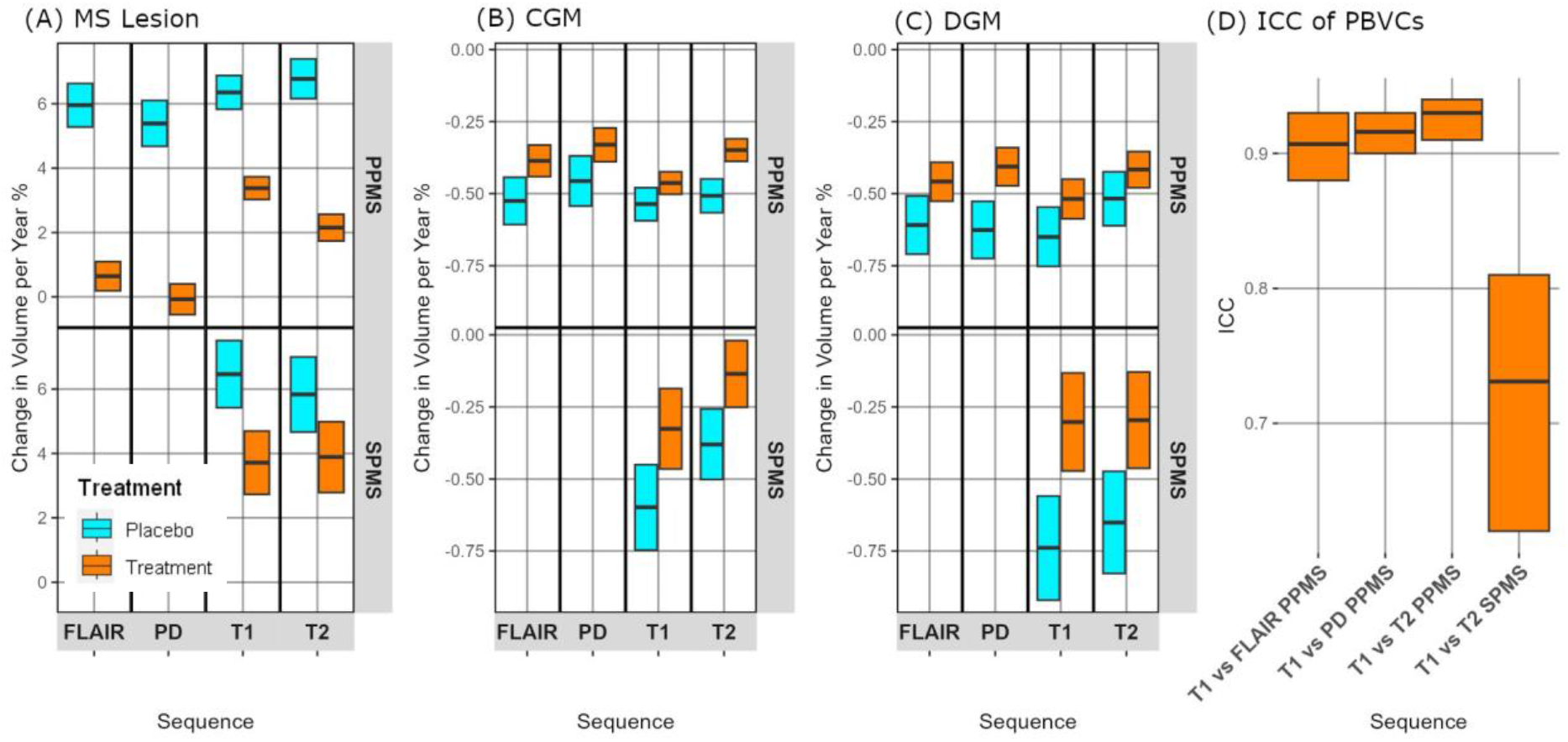
Measuring treatment effects using single MRI modalities. (A) illustrates the annual percent change in lesion volume detected by MindGlide across FLAIR, PD, T1, and T2 sequences for primary progressive MS (PPMS) and secondary progressive MS (SPMS) cohorts, stratified by treatment allocation. Notably, treatment cohorts exhibited a reduction in lesion volume accrual compared to placebo across all sequences. Boxes display medians and 95% CI. (B) depicts the annualized rate of cortical grey matter (CGM) atrophy. MindGlide successfully differentiated between treatment and placebo groups, demonstrating reduced cortical atrophy across all MRI sequences in treated patients. This is also the case for atrophy rates in deep grey matter (DGM) as seen in (C). There are no FLAIR and PD sequences available for the SPMS cohort. (D) shows inter-sequence reliability for percentage brain volume changes (PBVC): High intra-class correlation coefficients (ICC) for percent brain volume change (PBVC) across different MRI sequences, indicating high inter-sequence reliability. This underscores the segmentation tool’s robustness and consistency in detecting neurodegenerative changes across various imaging modalities. PPMS: N = 680, SPMS: N= 130

Figure 4 shows the rates of change across CGM, DGM and lesions in different MRI modalities in the real-world paediatric cohort. In T2 scans, lesion volumes increased by 0.612 ml/year in the moderate efficacy treatment group (P<0.001), while remaining stable in the high efficacy treatment group (model-estimated average -0.376 ml/year, P=0.230). FLAIR lesions showed stability in both groups: -0.063 ml/year in the high efficacy group (P=0.824) vs 0.009 ml/year in the moderate efficacy group (P=0.966). T1-weighted hypointense lesions remained stable in the high efficacy group (model-estimated average 0.165 ml/year, P=0.611) but increased by 0.647 ml/year in the moderate efficacy group (P=0.001).

**Figure 4.**
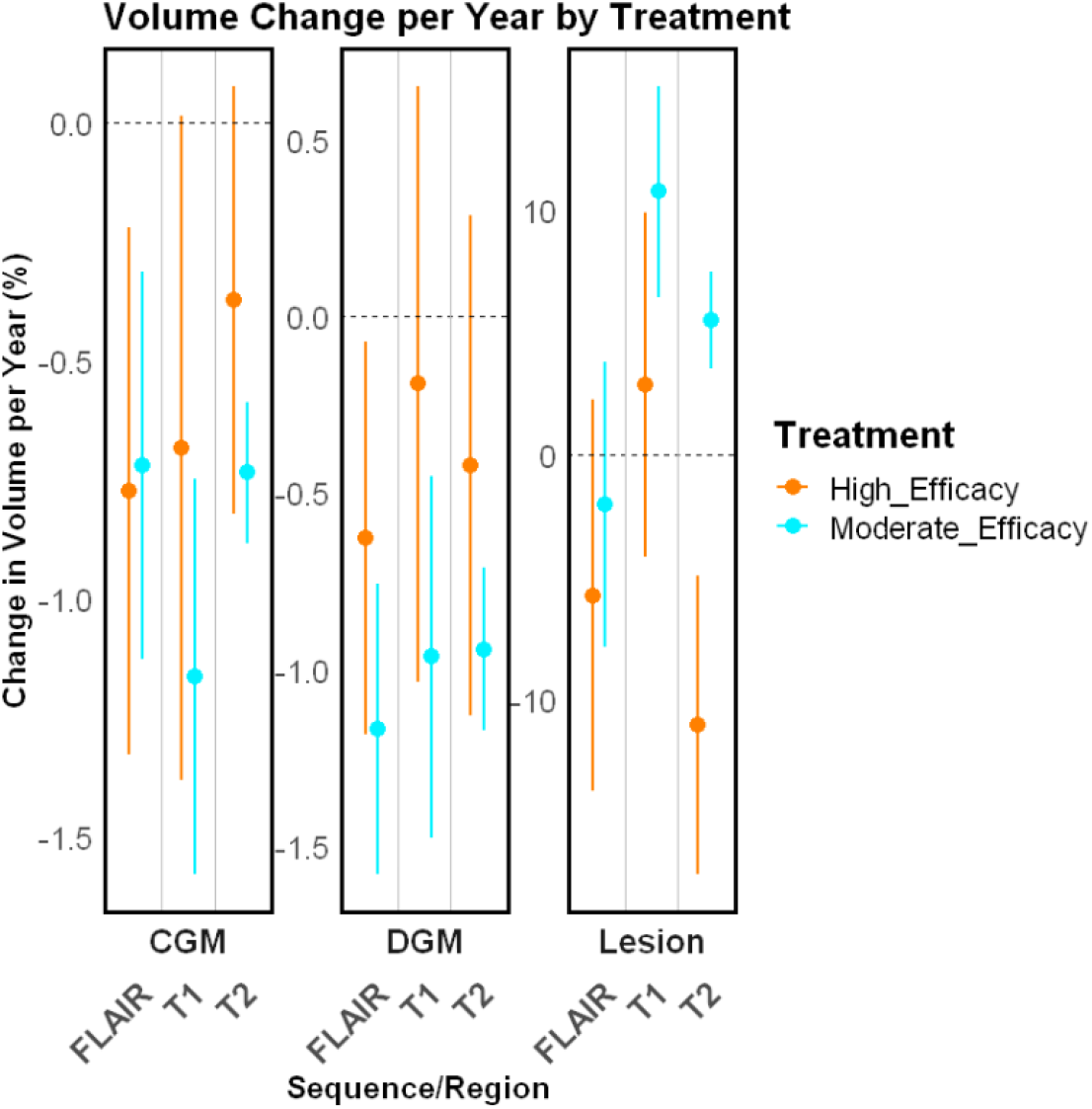
Longitudinal changes of brain regions and lesion volumes in the real-world paediatric dataset. Linear mixed-effects models for cortical grey matter, deep grey matter, and lesion volume on a paediatric real-world cohort, stratified by treatment allocation. Brain region volume changes over time in this real-world cohort. Median values are shown as a dot, and the whiskers show the 95% confidence intervals. N = 161. 72 patients received high-efficacy treatment, and 89 received moderate efficacy treatment. Abbreviations: FLAIR, Fluid Attenuated Inversion Recovery; CGM, Cortical Grey Matter; DGM, Deep Grey Matter.

### Treatment effects on brain atrophy

In the SPMS trial (simvastatin vs placebo), MindGlide demonstrated a significantly reduced rate of cortical GM volume loss in the treatment group across both T2-weighted and 3D T1-weighted MRI. Using T2-weighted MRI, the loss rate was 0.704 mL/year in the treatment group compared to a faster 1.792 mL/year in the placebo group (P=0.008). The 3D T1-weighted MRI analysis showed a similar treatment benefit, with a loss rate of 1.629 mL/year in the treatment group versus 2.912 mL/year in the placebo group (P=0.009).

When analysing DGM volume loss, MindGlide demonstrated a significantly slower loss rate in the treatment group than placebo across both T2-weighted and 3D-T1 weighted MRI modalities. With T2-weighted MRI, the loss rate was 0.101 ml/year in the treatment group versus 0.205 ml/year in the placebo group (P=0.009). We observed a similar treatment benefit in 3D-T1 weighted MRI analysis, showing a loss rate of 0.105 ml/year in the treatment group compared to 0.234 ml/year in the placebo group (P=0.001).

In the PPMS trial (ocrelizumab vs. placebo), MindGlide consistently demonstrated a slower cortical GM volume loss rate in the treatment group across different MRI modalities. Specifically, using T2-weighted MRI, the loss rate was 1.638 ml/year in the treatment group compared to 2.335 ml/year in the placebo group (P<0.001). We also observed treatment effect with T2-FLAIR (treatment: 1.778 ml/year vs. placebo: 2.342 ml/year, P=0.016), PD (treatment: 1.683 ml/year vs placebo: 2.310 ml/year, P=0.021), and showed a similar trend in 2D-T1 weighted MRI (treatment: 2.183 ml/year vs. placebo: 2.485 ml/year, P=0.06).

In the same PPMS trial, MindGlide analysis of DGM volume loss rate showed treatment effects depending on the MRI modality: T2-weighted imaging revealed no statistically significant difference (0.172 ml/year in the placebo group vs. 0.143 ml/year in the treatment group, P=0.130). However, treatment group analysis using T2-FLAIR (0.200 ml/year in the placebo group vs. 0.156 ml/year in the treatment group, p=0.028), PD (0.220 ml/year in the placebo group vs 0.144 ml/year in the treatment group, p<0.001), and T1-weighted MRI (0.212 ml/year in the placebo group vs. 0.172 ml/year in the treatment group, P=0.049) demonstrated a consistently slower loss rate. Figure 5 shows example segmentations across different modalities.

**Figure 5.**
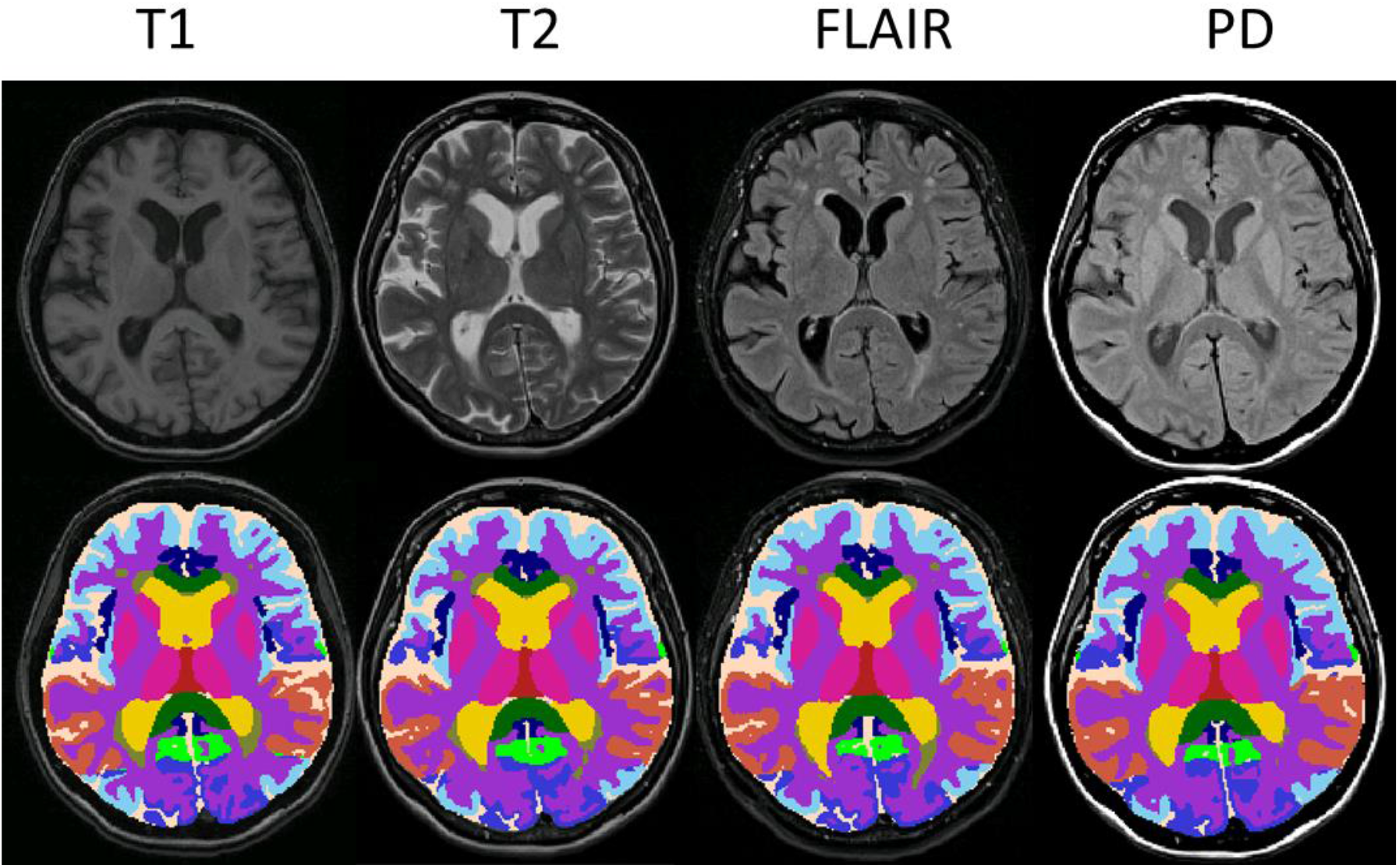
Example segmentations from various modalities. The figure shows separate segmentations by the MindGlide model on 2D T1-weighted, T2-weighted, FLAIR and PD modalities in the PPMS trial. The top row shows the unprocessed (“raw”) scans, and the bottom row shows labels or segmentations corresponding to anatomical regions, in addition to white matter hyperintensities (or hypo-intensities in the case of T1-weighted modality).

In the real-world paediatric cohort, we measured cortical grey matter loss in both treatment groups. In T1 images, we found a loss of 3.736 ml/year in the high efficacy group (P = 0.049) and 5.807 ml/year in the moderate efficacy group (P < 0.001). T2 images revealed a loss of 2.102 ml/year in the high efficacy group (P = 0.068) and 3.66 ml/year in the moderate efficacy group (P < 0.001). In FLAIR images, we observed a loss of 4.19 ml/year in the high efficacy group (P = 0.005) and 3.516 ml/year in the moderate efficacy group (P = 0.001).

The DGM volume for the real-world paediatric cohort stayed stable (model-estimated average -0.086 ml/year, P=0.552) in the high efficacy treatment group. Conversely, there was a volume loss of 0.301 ml/year in the moderate efficacy treatment group (P=0.001) in T1 images. In T2 scans there was no DGM volume change (model-estimated average -0.163 ml/year, P=0.207) in the high efficacy treatment group while there was a volume loss of 0.32 ml/year in the moderate efficacy treatment group (P<0.001). In FLAIR scans there was a volume loss of 0.22 ml/year (P=0.034) in the high efficacy treatment group and 0.399 ml/year (P<0.001) in the moderate efficacy treatment group.

### Reliability

#### Cross-sectional results

Figure 6 presents MindGlide’s label agreement across different MRI modalities of the same scans across 19 segmented regions. ICC values for brain regions ranged from 0.85 to 0.98, except for the optic chiasm (ICC 0.59). MS lesions demonstrated an ICC of 0.95 (95% CI [0.93, 0.95]) across modalities.

**Figure 6.**
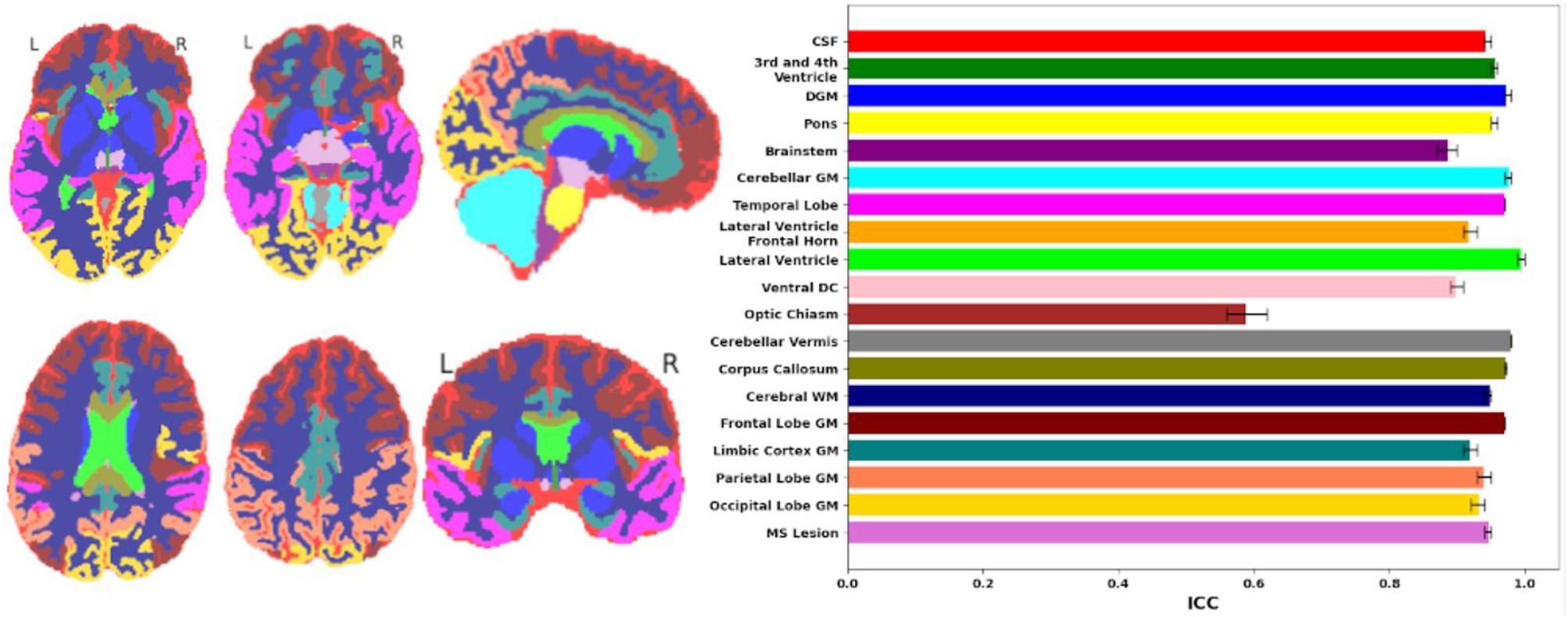
Reliability of regional segmentations across MRI modalities. Consistency of segmented regions or labels across multiple MRI modalities measured by the intraclass Correlation Coefficients (ICCs). On the left, coloured brain maps depict all 19 brain region labels: CSF (Cerebrospinal Fluid), 3^rd^ and 4^th^ Ventricle, DGM (Deep Grey Matter), Pons, Brainstem, Cerebellar GM (Grey Matter), Temporal Lobe, Lateral Ventricle Frontal Horn, Lateral Ventricle, Ventral DC (Diencephalon), Optic Chiasm, Cerebellar Vermis, Corpus Callosum, Cerebral WM (White Matter), Frontal Lobe GM, Limbic Cortex GM, Parietal Lobe GM, and Occipital Lobe GM, along with MS (Multiple Sclerosis) Lesions. The right side presents ICC values ranging from 0 to 1 for these regions, providing a quantitative measure of the consistency across multiple MRI sequences. Higher ICC values indicate greater reliability in the measurement of a particular brain region. N = 699.

#### Longitudinal: Inter-Sequence Consistency of Brain Atrophy

In addition to Dice score comparisons across sequences, we evaluated the comparability between longitudinal changes in different MRI modalities. In the PPMS dataset, T1 vs. FLAIR showed an ICC of 0.91 (95% CI [0.88, 0.93]), T1 vs PD had an ICC of 0.916 (95% CI [0.90, 0.93]) and T1 vs. T2 we calculated an ICC of 0.93 (95% CI [0.91, 0.94]). In the SPMS dataset, the ICC between T1 and T2 was 0.73 (95% CI [0.62, 0.81]). All the images used for this analysis were 2D. This suggests that PBVC, as a surrogate measure of neurodegeneration, is reliably assessed by our segmentation tool irrespective of the MRI sequence employed.

## DISCUSSION

Our work establishes the capability to extract multiple clinically relevant MS biomarkers from a single MRI modality. This advancement significantly streamlines analysis and opens the door to large-scale research using diverse and often incomplete clinical MRI datasets – a significant advantage for clinical trials and real-world studies. MindGlide demonstrates superior performance to existing tools in correlating biomarkers with disability. Furthermore, it accurately captured treatment effects on disease activity (as shown by lesion accrual) and neurodegeneration (as shown by brain tissue loss) across clinical trials.

Our results demonstrate that meaningful tissue segmentation and lesion quantification are achievable even with limited MRI data and single modalities not typically used for these tasks (e.g., T2-weighted MRI without FLAIR). We established the validity and reliability of these findings both cross-sectionally and longitudinally. At baseline, volumetric measurements extracted using our method showed a higher correlation with EDSS than an established modality-agnostic segmentation method (SAMSEG), demonstrating the validity of our approach. Longitudinally, MindGlide-derived brain volume measurements revealed reduced atrophy in treated groups in two unseen clinical trials. This finding was consistent across MRI sequences when scanners and protocols remained unchanged over time.

Our work contributes to the evolution of MRI processing tools that streamline previously time-consuming pipelines. Running the model on consumer-grade GPU hardware took, on average, 37 seconds. Efficiency is especially valuable for MS MRIs, where multimodal imaging is traditionally used to extract biomarkers. The typical workflow has involved intensity inhomogeneity correction^33^, followed by automatic segmentation of white matter lesions using T2-FLAIR and three-dimensional T1-weighted MRI. To mitigate the misclassification of hypointense lesions as grey matter (which share a similar intensity profile), anatomical T1-weighted MRIs may undergo lesion filling after affine registration with T2-FLAIR images^34^. Subsequently, hand-labelled T1-weighted MRIs (known as atlases) are non-linearly registered to the subject’s T1-weighted MRI ^35,36^. Labels from the co-registered atlases are then fused using various fusion algorithms, followed by probabilistic segmentation to differentiate tissue classes (white matter, grey matter, and cerebrospinal fluid) ^37^. Moving towards newer deep-learning-based pipelines allows fast turnaround times for automatic MRI analysis and real-world implementations.

Our study benefits from a large sample size for model training and external validation. Importantly, our findings generalised across datasets and MRI modalities. Our training used only FLAIR and T1 images, yet the model successfully processed new modalities (like PD and T2) from different scanners and periods encountered during external validation. This success is due to the SynthSeg algorithm’s domain randomisation during synthetic data generation (or augmentation), enabling cross-modality generalisation as has been shown before^7,13^.

While we found high ICC values across all segmented brain structures, interpreting the ICC values for MS lesions warrants a more nuanced consideration. We analysed the ICC across different MRI sequences, including FLAIR, T2, T1, and PD. We analysed the ICC across different MRI sequences during external validation, including FLAIR, T2, T1, and PD. While the MindGlide model was trained on both T1 and FLAIR images, the lesions used for training came from the FLAIR sequence but the domain randomisation enabled the model to generalise to unseen modalities. While improving generalisability, this approach blurs pathological specificity. T1 hypointensities can be pathologically distinct from FLAIR hyperintensities. Therefore, while the ICC values provide an essential insight into the tool’s reliability, the difference in pathophysiological representation between sequences necessitates a careful approach to interpreting these results. For example, chronic T1 hypo-intensities, often called “black holes,” indicate MS lesions characterised by axonal loss and tissue destruction^38^. The ICC values obtained for MS lesions must, therefore, be considered within the context of these different imaging signatures. A high ICC value might suggest that while the segmentation tool is consistent across sequences, it may not fully distinguish the complex nature of lesion pathology that varies between T1 and T2/FLAIR sequences. Therefore, the lack of pathological specificity of modality-agnostic models remains a limitation. Nonetheless, as explained above, this approach enables using data to enable a new avenue of research on archival real-world, data.

Overall, segmenting different structures was highly reliable except for optic chiasm, which had moderate reliability (ICC of 0.59). This can be explained by the smaller size of the optic chiasm compared to all other MindGlide labels. A single voxel discrepancy within this region wields a proportionately larger impact on the ICC, magnifying the effect of any spatial variations. Furthermore, its close encirclement by cerebrospinal fluid (CSF) can obscure the chiasm’s boundaries in imaging sequences, reducing the dice score.^39^ The intra-class correlation analysis across different MRI sequences demonstrated the consistency and reliability of PBVC measurements obtained by MindGlide. We chose the number of labels or segmentations to be 19 by merging smaller labels from the atlas to ensure efficiency and lower computational expense during inference time with a view for under-resourced research settings (e.g., hospitals). Therefore, current implementation is not intended for detailed segmentations (for example a thalamic volume instead of deep grey matter volume) of brain structures.

Future MindGlide development will focus on specialised fine-tuned models for MRI sequences to provide more pathological specificity in real-world data. For example, a new class of models can be fine-tuned for detecting T1-hypointensities and slowly evolving lesions^40^. Or incorporating gadolinium-enhancing lesions would help differentiate active MS inflammation^41^. Additionally future will expand MindGlide with self-supervised learning to reduce the need for manual labels, and implement it across several UK hospitals for real world research.

## Data Availability

All data produced in the present study are available upon reasonable request to the authors

## Acknowledgements

This study/project is funded by the National Institute for Health and Care Research (NIHR) Advanced Fellowship Round 7 to Dr Arman Eshaghi (NIHR302495). The views expressed are those of the author(s) and not necessarily those of the NIHR or the Department of Health and Social Care.

## Supplemental Methods

### Model architecture

The MindGlide model is a 3D-CNN model using hyperparameters obtained from a recent international challenge of the nnU-net model and implemented using the “dynamic Unet architecture (DynUNet)” inside the MONAI framework^11,12,29^.

MindGlide takes 3D images with one input channel and outputs 20 segmented channels, each representing a unique label, including the background. The model is structured into several sections: an input block, a series of down-sampling blocks, a series of up-sampling blocks, an output block, multiple deep supervision heads, and skip layers to facilitate information flow across the network. The input block consists of two convolutional layers (Conv3d) with Leaky ReLU activations and Instance Normalization^42^. The down-sampling section consists of four “Unet basic block” units, each composed of two 3D convolution layers with instance normalisation and LeakyReLU activation. The bottleneck again comprises two convolution layers with instance normalisation and LeakyReLU activation, performing the core extraction of features. The up-sampling section consists of five “Unet up block” units. Each upsample block consists of a ConvTranspose3d layer for up-sampling, followed by a basic Unet block, which contains two Conv3d layers with instance normalisation and LeakyReLU activation. The output block consists of a single 3D convolution layer.

The deep supervision heads consist of three convolutional layers, each one performing convolutions to gradually reduce the number of feature channels to the desired number of output channels (20 in our case). This design comprises 43 layers with 30,781,744 trainable parameters. See the Code Availability section for the URL to our code, pre-trained models and the MONAI implementation (https://www.monai.io).^29^

### Evaluating MindGlide with “ground truth” manual lesion segmentations cross sectionally and longitudinally

We used two openly available lesion segmentation datasets as ground truth comparators^30,31^. The first dataset, MS-30, had 30 MS patients (23 women) with median age 39 years (range 25-64) and relapse-onset MS^31^. Each patient had pre-processed and co-registered 3D FLAIR, T1-weighted, and T2-weighted images, and a consensus lesion mask generated by three raters (details in ^31^). The second dataset, ISBI, was from the 2015 International Symposium on Biomedical Imaging Lesion Challenge^43^. It had a training set of 5 patients, each with four time-separated scans for longitudinal lesion segmentation. Each patient and timepoint had co-registered FLAIR, T1, T2 and proton density data. Two independent raters provided lesion masks, but no consensus mask. We used the most experienced rater (17 years) for cross-sectional analysis. We restricted analysis to FLAIR lesion segmentations and ran MindGlide and SAMSEG on all FLAIR images. For cross-sectional lesion segmentation, we pooled the datasets to create n=50 FLAIR lesion image masks from 35 patients. We assessed lesion load (mm3), voxel-wise spatial metrics (Dice overlap scores, sensitivity, precision) to compare segmentation software against reference masks.

To assess lesion segmentation longitudinally, we used the ISBI dataset to calculate the Intraclass Coefficient (ICC) between rater 1 and rater 2, rater 1 and MindGlide and for completeness, rater 2 and MindGlide. To this end we performed a linear mixed effects model with lesion volume as the dependent variable, rater as a fixed effect and both time and subject as random effects (R software, lme4 package). The ICC was calculated from the variance of the model output and defined as the total rater variance divided by the total rater variance + the residual variance of the model.

## Supplemental Results

### Lesion Segmentation Analysis

Lesion load was calculated from each software and reference rater mask. Mean lesion loads were 15456.6 ± 14399.9 mm^3^ for the ground truth, 14250.3 ± 12447.3 mm^3^ for MindGlide and 6296.4 ± 6331.2 mm^3^ for SAMSEG. One-way ANOVA revealed a significant Group effect (F(_1,48)_ = 9.234, *p* < 0.001) and post hoc pairwise comparisons revealed a significant difference between lesion load calculated from ground truth masks vs SAMSEG (p < 0.001) and MindGlide vs SAMSEG (p < 0.01) (see Figure 3 in the main text) with SAMSEG defining smaller lesions compared to both ground truth masks and MindGlide lesion segmentations.

All spatial metric summary data and statistics are presented in Table 2.

**Table 2.** Treatment effects in the external validation clinical trials across different MRI modalities.

| **Trial** | **Variable** | **Sequence<sup>1</sup>** | **Placebo**<br>**Volume**<br>**Change**<br>[95% CI] | **Treatment**<br>**Volume**<br>**Change**<br>[95% CI] | **p-value<sup>2</sup>** |
| --- | --- | --- | --- | --- | --- |
| SPMS<br>(simvastatin<br>vs placebo) | Hyperintense<br>lesions | T2 | 1.122<br>[0.987, 1.256] | 0.768<br>[0.520, 1.017] | 0.055 |
|  | Hypointense<br>lesions | 3D-T1 | 1.875<br>[1.666, 2.084] | 1.071<br>[0.686, 1.457] | 0.006 |
|  | CGM | T2 | -1.792<br>[-2.089, -1.495] | -0.704<br>[-1.254, 0.155] | 0.008 |
|  |  | 3D-T1 | -2.912<br>[-3.266, -2.558] | -1.630<br>[-2.283, -0.976] | 0.009 |
|  | DGM | T2 | -0.205<br>[-0.234, -0.176] | -0.102<br>[-0.155, -0.048] | 0.009 |
|  |  | 3D-T1 | -0.234<br>[-0.263, -0.204] | -0.105<br>[-0.159, -0.050] | 0.002 |
| PPMS<br>(ocrelizumab<br>vs placebo) | Hyper<br>intense<br>Lesions | T2-FLAIR | 1.042<br>[0.911, 1.174] | 0.141<br>[0.053, 0.229] | <0.001 |
|  | Hyper<br>intense<br>Lesions | PD | 0.633<br>[0.535, 0.731] | 0.091<br>[0.025, 0.157] | <0.001 |
|  | Hypointense<br>lesions | T1 | 1.225<br>[1.112, 1.338] | 0.649<br>[0.573, 0.725] | <0.001 |
|  | Hyper<br>intense<br>Lesions | T2 | 1.104<br>[0.990, 1.217] | 0.400<br>[0.324, 0.476] | <0.001 |
|  | CGM | T2-FLAIR | -2.342<br>[-2.722, -1.963] | -1.778<br>[-2.033, -1.524] | 0.016 |
|  |  | PD | -2.310<br>[-2.752, -1.868] | -1.683<br>[-1.980, -1.386] | 0.021 |
|  |  | T1 | -2.485<br>[-2.753, -2.217] | -2.183<br>[-2.363, -2.002] | 0.066 |
|  |  | T2 | -2.335<br>[-2.606, -2.065] | -1.638<br>[-1.820, -1.457] | <0.001 |
|  | DGM | T2-FLAIR | -0.200<br>[-0.234, -0.167] | -0.156<br>[-0.178, -0.133] | 0.028 |
|  |  | PD | -0.220<br>[-0.255, -0.186] | -0.144<br>[-0.167, -0.121] | <0.001 |
|  |  | T1 | -0.212<br>[-0.245, -0.179] | -0.172<br>[-0.194, -0.150] | 0.049 |
|  |  | T2 | -0.172<br>[-0.203, -0.141] | -0.143<br>[-0.164, -0.123] | 0.130 |

<sup>1</sup>All the MRI sequences were two dimensional or 2D unless specified as 3D (1x1x1 mm). Volume changes are in ml. The real-world cohort is not included here because there was no untreated group (41% received Interferon Beta, 38% Ocrelizumab and the rest other disease modifying treatments).
<sup>2</sup>treatment effect p value, or p value of the difference between volume change rates in treatment vs placebo groups from the mixed effects model.
Abbreviations: 95% CI, 95% confidence interval; CGM, cortical grey matter; DGM, deep grey matter; FLAIR, Fluid-Attenuated Inversion Recovery; PD, proton density; 2D, two dimensional (non-isotropic voxels); 3D, three dimensional (isotropic 1x1x1mm resolution); SPMS, secondary progressive multiple sclerosis; PPMS, primary progressive multiple sclerosis.

An ICC analysis assessed how well MindGlide performed at segmenting lesions from the same patient across multiple time points. For this, we calculated ICC agreement values for rater 1 vs rater 2, rater 1 vs MindGlide and rater 2 vs MindGlide, with the ICC values being 0.98, 0.97 and 0.96, respectively.

### Treatment Effect Analysis on Percentage Brain Volume Change

We calculated treatment effects on percentage brain volume change (PBVC) for every MRI modality in our PPMS dataset (Supplemental Figure 1). Annualised PBVC in T1-weighted images was -0.91% in the placebo group and -0.85% in the treatment group (p-value = 0.572). In T2-weighted images PBVC was -0.65% in the placebo group and -0.56% in the treatment group. (p-value = 0.279). In FLAIR images PBVC was -1.22% in the placebo group and -1.12% in the treatment group (p-value = 0.442) and in PD images PBVC was -0.95% in the placebo group and -0.81% in the treatment group, respectively (p-value = 0.232). Despite showing slower brain volume loss over time in the treatment groups across all MRI modalities, all the results were statistically not significant.

## Supplemental Tables

**Supplemental Table 1.** Data sources used for training and external testing of MindGlide.

| **Trial** | **Included in this study\*** |
| --- | --- |
| **Training datasets** |  |
| ADVANCE <sup>14</sup> | 959 |
| ASCEND <sup>15</sup> | 997 |
| BRAVO <sup>16</sup> | 494 |
| DCE (DEFINE,<br>CONFIRM, ENDORSE)<br><sup>17–19</sup> | 53 |
| EXPAND <sup>3</sup> | 1004 |
| INFORMS <sup>21</sup> | 344 |
| SPI2 <sup>22</sup> | 373 |
| Observational UCL<br>cohort (PITMS cohort,<br>adults with MS) | 23 |
| **External validation** |  |
| Paediatric MS | 161 |
| ORATORIO<sup>2</sup> | 699 |
| MS-STAT <sup>23</sup> | 141 |

\* We sampled the training data from the International Progressive MS Alliance data repository at the Montreal Neurological Institute.

**Supplemental Table 2.**
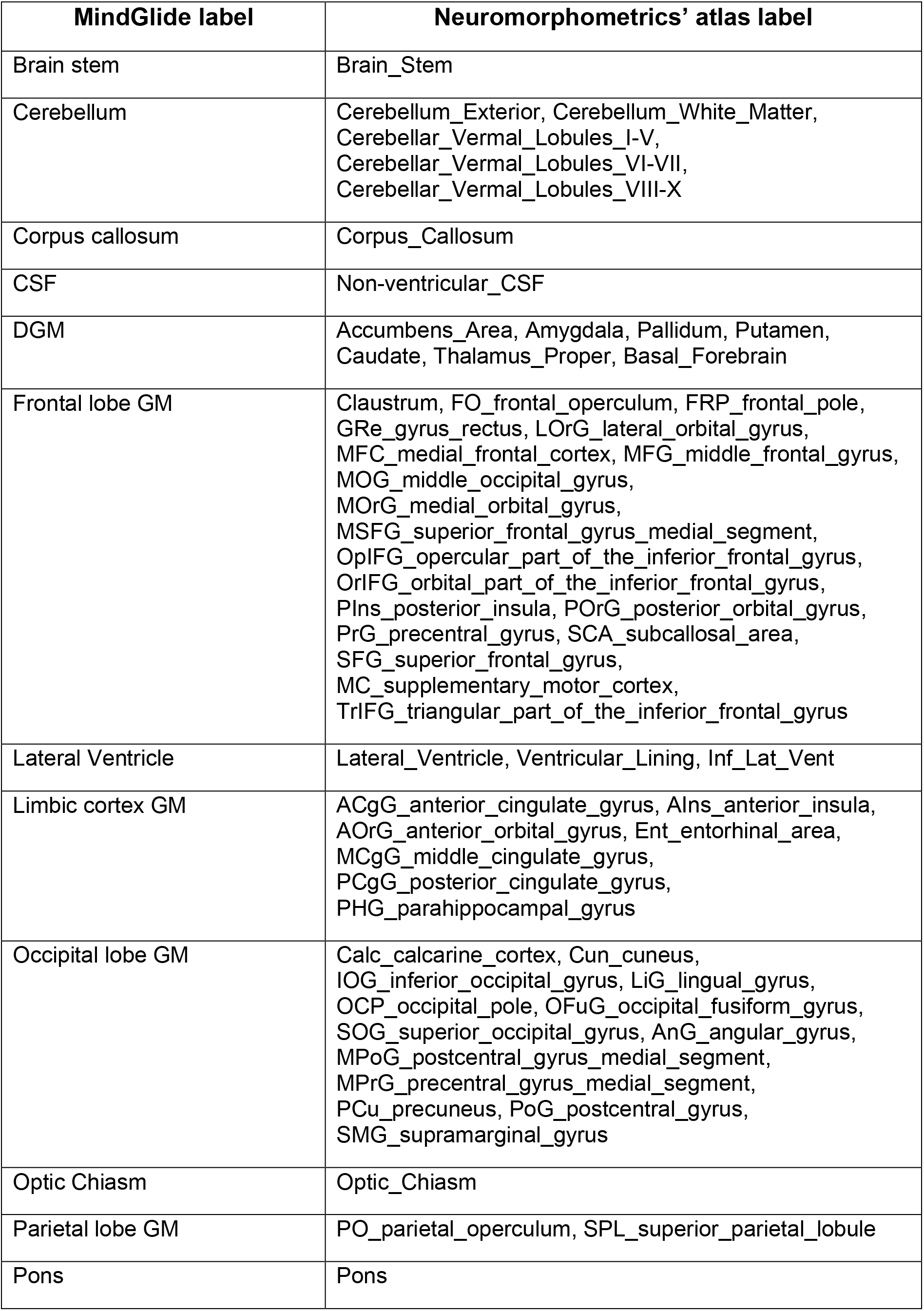
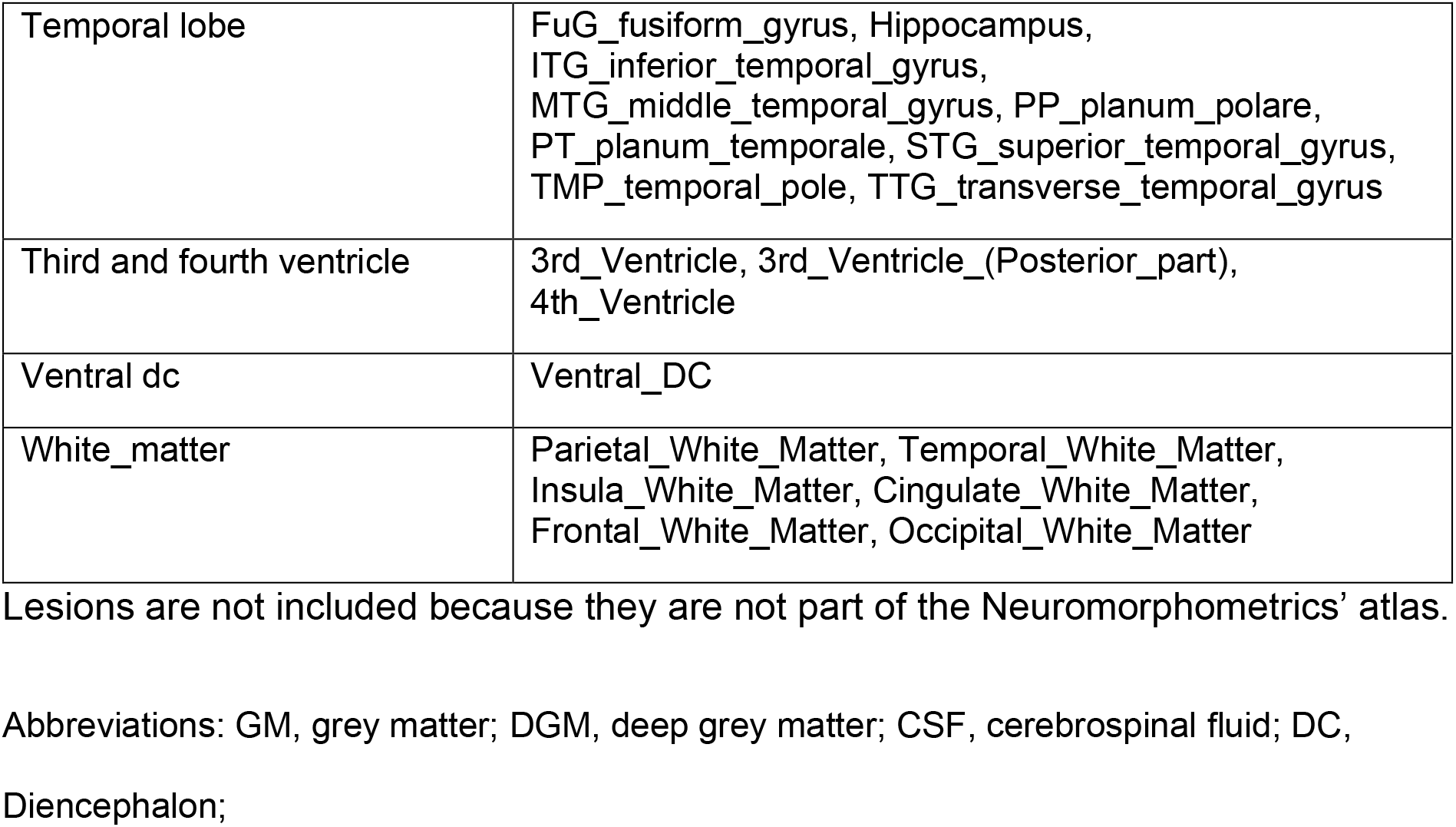
Correspondence between MindGlide labels and Neuromorphometrics’ atlas.

**Supplemental Table 3.**
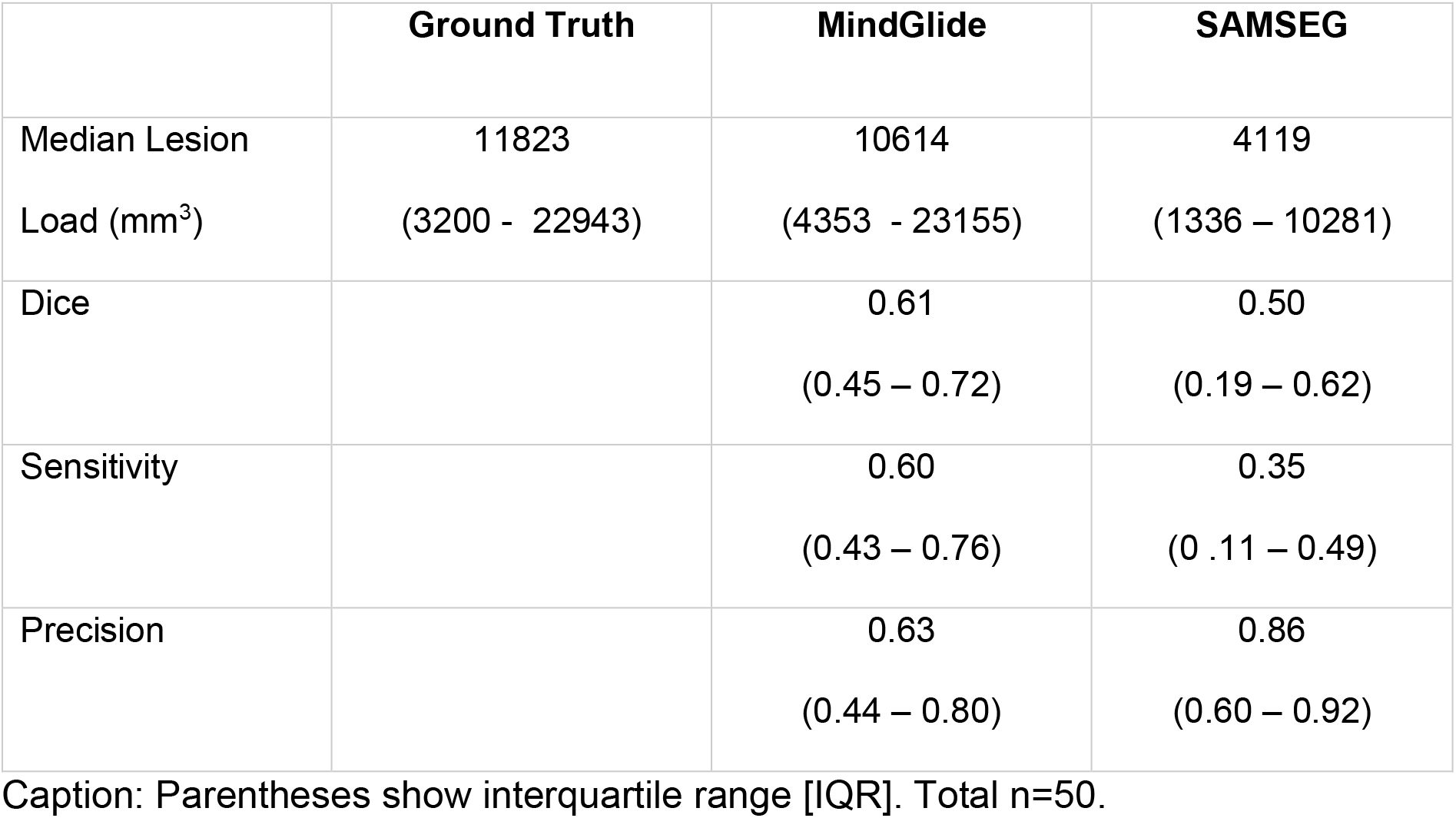
Cross-software comparison of lesion segmentation (MindGlide and SAMSEG).

**Supplemental Table 4:** Median running time of a MindGlide model on a single MRI modality during inference.

| **Model** | **Runtime**<br>**(IQR)** | **GPU** | **GPU**<br>**Memory** | **Maximum GPU**<br>**Memory used during**<br>**Inference** |
| --- | --- | --- | --- | --- |
| Mindglide | 37 seconds<br>(34 - 40) | NVIDIA Quadro RTX<br>6000 | 24 GB | 7850 MiB |

Runtime was analyzed across 10109 scans and maximum used GPU memory across 90 scans.

## Supplemental Figures

**Supplemental Figure 1.**
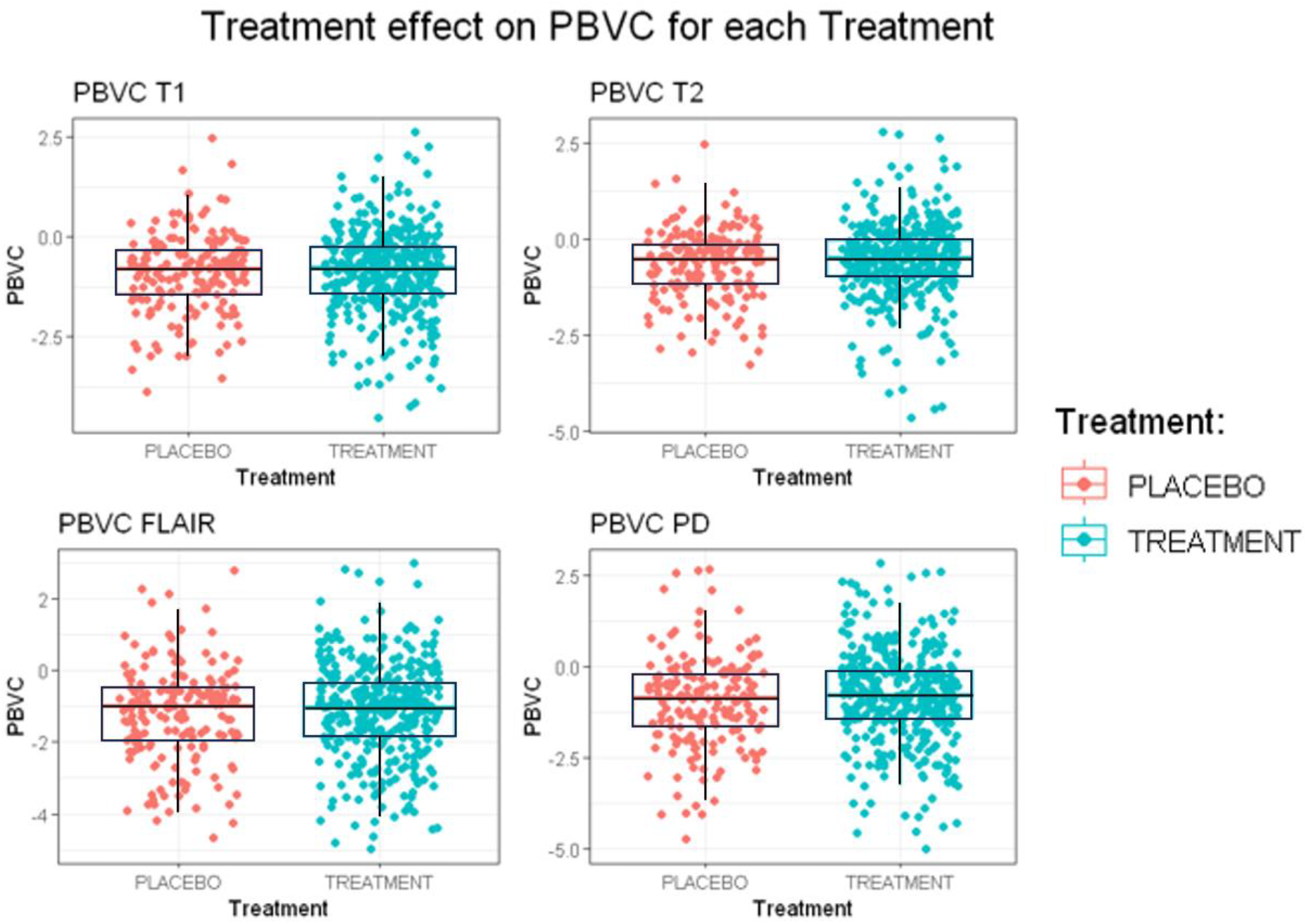
Brain atrophy in treated and control arms. Supplemental Figure 1 shows boxplots and individual data points, illustrating the distribution and variability of PBVC per year measurements across four different MRI sequences in the PPMS dataset: T1-weighted, T2-weighted, FLAIR, and Proton Density (PD). The data is categorized by two treatment groups, to evaluate the treatment effect on PBVC. Red points indicate the placebo group, while turquoise points represent the treatment group. We found less atrophy in the treatment group compared to the placebo group in every sequence. However, none of these were statistically significant. To enhance the graph’s visual clarity, 14 data points classified as outliers have been intentionally omitted from the visualization. Only patients with a follow-up of at least 2 years were used. N = 576.

### Pairwise comparison of PBVCs of each sequence

**Supplemental Figure 2.**
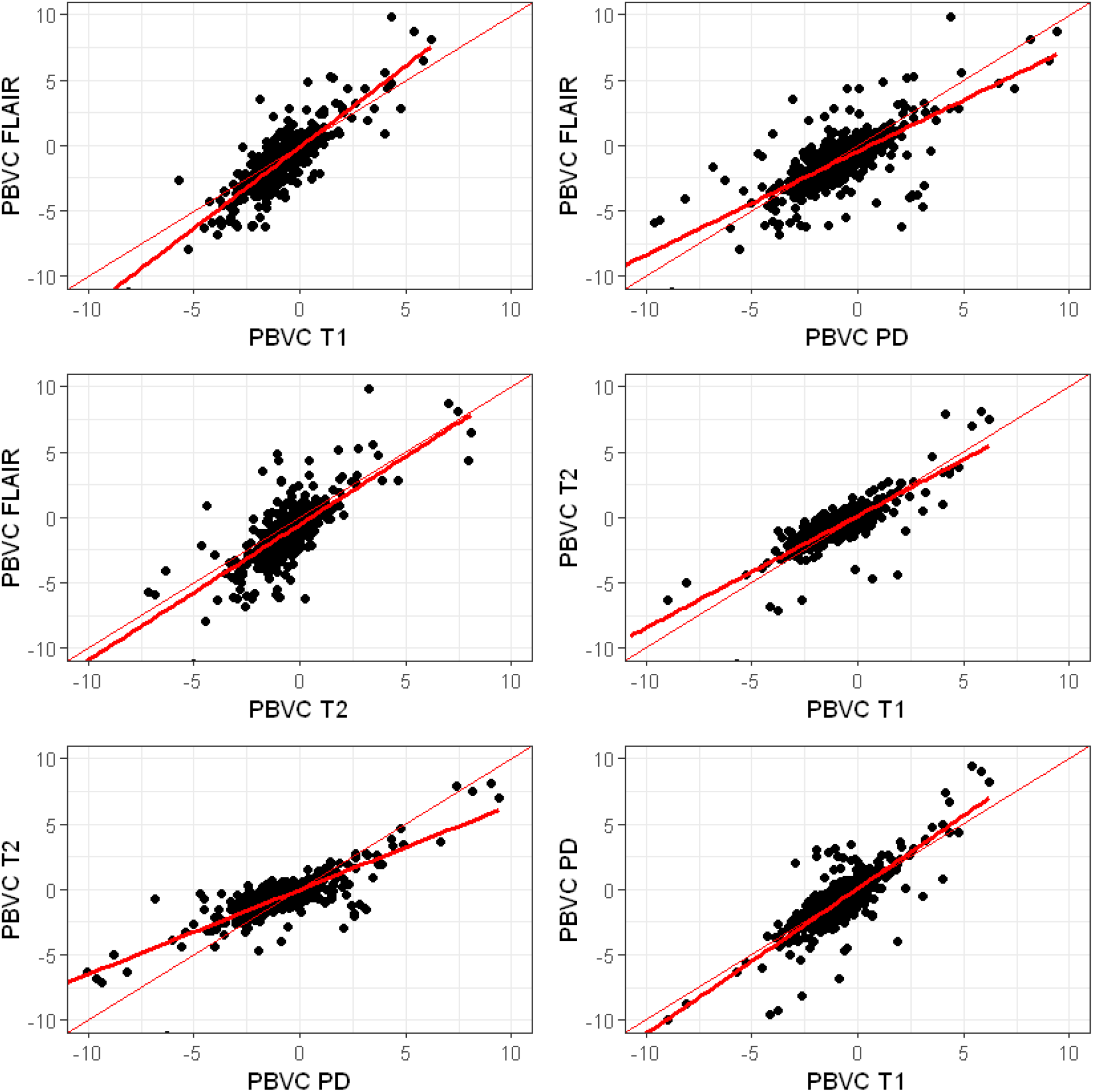
Scatter Plots of PBVC Measurements across different MRI Sequences. Supplemental Figure 2 shows a series of scatter plots to visualise the pairwise comparisons of Percent Brain Volume Change (PBVC) per year measured from different MRI sequences in the PPMS dataset: T1-weighted, T2-weighted, Proton Density (PD), and FLAIR. Each plot displays the relationship between two distinct MRI sequence measurements, highlighted with points representing individual observations. The thinner line in each plot represents a line of unity (slope = 1, intercept = 0), serving as a reference for perfect agreement between the measurements from the two MRI sequences. Linear regression lines (the thicker red line) indicated the regression line fitted to data. N = 680

## Disclosures

In the past three years, Arman Eshaghi has received research grants from the Medical Research Council (MRC), National Institute for Health and Social Care Research (NIHR), Innovate UK, Biogen, Merck, and Roche. He has served as an advisory board member of Merck Serono and Bristol Myers Squib. He is the founder and equity stakeholder in Queen Square Analytics Limited. He serves on the editorial board of Neurology (American Academy of Neurology).

## Notes

### Competing Interest Statement

The authors have declared no competing interest.

